# Clinical evaluation of the Roche/SD Biosensor rapid antigen test with symptomatic, non-hospitalized patients in a municipal health service drive-through testing site

**DOI:** 10.1101/2020.11.18.20234104

**Authors:** Zsὁfia Iglὁi, Jans Velzing, Janko van Beek, David van de Vijver, Georgina Aron, Roel Ensing, Kimberley Benschop, Wanda Han, Timo Boelsums, Marion Koopmans, Corine Geurtsvankessel, Richard Molenkamp

**Affiliations:** Department of Viroscience, Erasmus MC, Rotterdam, The Netherlands; Public Health Service Rotterdam-Rijnmond, Rotterdam, The Netherlands; Centre for infectious Disease Control (Cib), National Public Health Institute (RIVM), Bilthoven, The Netherlands

**Keywords:** SARS-CoV-2, rapid antigen test, clinical evaluation, diagnostics

## Abstract

**Background:** Rapid detection of infectious individuals is essential in stopping the further spread of SARS-CoV-2. Although rapid antigen test is not as sensitive as the gold standard RT-PCR, the time to result is decreased by day(s), strengthening the effectiveness of contact tracing.

**Methods:** The Roche/SD Biosensor lateral flow antigen rapid test was evaluated in a mild symptomatic population at a large drive through testing site. A second nasopharyngeal swab was directly tested with the rapid test on site and results were compared to RT-PCR and virus culture. Date of onset and symptoms were analysed using data from a clinical questionnaire.

**Results:** We included 970 persons with complete data. Overall sensitivity and specificity were 84.9% (CI95% 79.1-89.4) and 99.5% (CI95% 98.7-99.8) which translated into a positive predictive value of 97.5% (CI95% 94.0-99.5) under the current regional PCR positivity of 19.2%. Sensitivity for people with high loads of viral RNA (ct <30, 2.17E+05 E gene copy/ml) and who presented within 7 days since symptom onset increased to 95.8% (CI95% 90.5-98.2). Band intensity and time to result correlated strongly with viral load thus strong positive bands could be read before the recommended time. Around 98% of all viable specimen with ct <30 were detected successfully indicating that the large majority of infectious people can be captured with this test.

**Conclusion:** Antigen rapid tests can detect mildly symptomatic cases in the early phase of disease thereby identifying the most infectious individuals. Using this assay can have a significant value in the speed and effectiveness of SARS-CoV-2 outbreak management.

**Summary:**

- People with early onset and high viral load were detected with 98.2% sensitivity.
- 97% of individuals in which virus could be cultured were detected by the rapid test.
- This test is suitable to detect mild symptomatic cases.

## Introduction

The Severe Acute Respiratory Syndrome Coronavirus 2 (SARS-CoV-2) has emerged almost a year ago [1] but it still keeps a strong grip not only on our daily life but also on the diagnostic capacities. Reverse transcriptase polymerase chain reaction (RT-PCR) has been the gold standard for diagnosis of acute infection [2] but has several limitations, such as the requirement for specialised laboratory infrastructure, trained personnel and reagents that have been in shortage globally [3]. In addition, the current turnaround time from sample collection to reporting of the result may take up to more than 48 hours [4] mainly due to the need to transport samples to laboratories, compromising effectiveness of triage, isolation and contact tracing strategies.

Rapid antigen detection tests (Ag RDT) for SARS-CoV2 appeared on the market earlier 2020 but initial reports of poor performance and the lack of independent evaluation results made governments reluctant to invest and consider the inclusion into testing algorithms. Currently there are 73 (and the number is growing) assays on the market [5] but few have been extensively validated [5-7]. Initial results showing that these tests are suitable detecting early onset cases with high viral load. As expected, the sensitivity of the tests is lower compared to RT-PCR, but in patients in the early phase of illness onset and with high viral load the performance meets the WHO set criteria of at least ≥80% sensitivity and ≥97% sensitivity compared to nucleic acid detection methods as gold standard [8]. Thus these tests could be useful in identifying the most infectious individuals [4]. In an outbreak scenario diagnostics with lower sensitivity but a faster result can render interventions more effective than gold standard tests [9]. Implementation of Ag RDT into testing algorithms would allow rapid detection and isolation of new cases and thereby support the test, trace and isolate strategy, aiming to stop transmission chains and reduce the impact of COVID-19.

In this study we aimed to assess the performance of the SD Biosensor SARS-CoV-2 Rapid Antigen Test (distributed by Roche Diagnostics) compared to both RT-PCR and virus culture in 970 individual. The field evaluation study was carried out at a large municipal health facility where people presented mostly with mild symptoms. Every individual over 18 years of age with an appointment for SARS-CoV-2 RT-PCR testing was approached to be included. An additional nasopharyngeal swab was obtained for the Ag RDT in parallel and processed on-site in order to compare sensitivity/specificity to RT-PCR. All Ag RDT and PCR positive samples were cultured to correlate results with infectivity.

## Methods

### Testing population, setup and patient recruitment

The study was carried out at the largest drive through testing location in Rotterdam - Rijnmond (the second largest city in the Netherlands) which is by appointment only. Eligibility for a free of charge test includes either symptoms or close contact with a confirmed SARS-CoV-2 infected person. Most people presenting for testing had (mild) symptoms. At the entrance of the testing site all people over 18 years of age were approached for inclusion and following informed consent people were enrolled and directed to one of the dedicated testing stations for the Ag RDT. The people that were included were also asked to fill in a clinical questionnaire including the reason for appointment, date of onset or end date and a list of symptoms (fever, sore throat, coughing, shortness of breath / tightness, runny nose, diarrhoea, eye complaints, nausea, rash, chills, headache, pain when breathing, coughing phlegm, muscle pain, painful / swollen lymph nodes, fatigue, vomiting, joint pain, loss of appetite, nosebleed, other). The study was carried out for 5 days in order to achieve the target of 800-1000 inclusions.

### Testing site setup and the mobile laboratory

From the six available testing posts we designated two posts for sample collection from people who enrolled in this study. This was based on: i) the average number of study subjects per test post (approximately 150 per day), ii) the known number of appointments per day and iii) the expected enrolment rate based on initial results from other study sites within the Netherlands. We expected to include a maximum of 300 people per day. Swabbing was done by the regular crew of trained personnel to avoid variations to the process. Testing was done on a benchtop, in a mobile unit (kindly provided by RIVM) by trained staff dressed in full personal protective equipment (goggles, FFP3 mask, gloves and disposable gown; PPE). Samples for the Ag RDT were collected at regular intervals and processed in convenient batches (5-10 tests at one time). Training, when necessary, was given on site and did not take longer than 30 mins. Following readout results were recorded in an offline database (Microsoft Access) designated for this study and both the swab and RDT device were inactivated in chlorine and disposed of as biohazard material.

### Specimen collection, testing and culture procedures

Standard method for SARS-CoV-2 testing is by RT-PCR which was carried out as usual, in parallel with the Ag RDT. Two swabs (one oro- and one nasopharyngeal swab, OP and NP swab) were taken for RT-PCR and virus culture, placed directly in universal transport media (HiViral™) and shipped to the Erasmus MC Viroscience diagnostic laboratory. For the Ag RDT evaluation a second nasopharyngeal swab was taken from the same nostril using the swab included in the kits to directly compare RT-PCR result with the Ag RDT. Routine RT-PCR testing was performed on combined oro- and one nasopharyngeal swab in virus transport medium using the cobas ® SARS-CoV-2 test on the COBAS6800 (Roche diagnostics). Genome copies/ml were calculated based on an in house established standard curve. The virus transport medium from the same oro- and nasopharyngeal swabs were also directly inoculated onto Vero cells clone 118, without prior freezing [10]. Samples were cultured for seven days, and, once cytopathic effect (CPE) was visible, the presence of SARS-CoV-2 was confirmed with immunofluorescent detection of SARS CoV-2 nucleocapsid protein (Rabbit polyclonal antibody Sino Biological inc.).

For the Ag RDT the SARS-CoV-2 Rapid Antigen Test (Distributed by Roche (SD Biosensor) (REF No. 9901-NCOV-01G; LOT No QCO3020079/Sub:A-2) was carried out immediately on-site following manufacturer’s instructions. A 4 grade scaling readout was utilized representing the strength of the band (++; +; +/-, -) and time till positive results was logged as less than 5 mins, less than 10 mins or 15 mins. When results were dubious, readout was performed independently by two persons.

### Data analysis

Data from the Ag RDT, RT-PCR, virus culture, and clinical questionnaire was merged using Microsoft Access, and data analysis was performed using R version 4.0.2. Sensitivity and specificity of Ag RDT was calculated in relation to the RT-PCR results as the gold standard. The Wilcoxon score interval was used to determine confidence intervals of proportions.

### Ethical clearance

The medical research ethics committee (MREC) of Utrecht decided the study was not subject to the Medical Research Involving Human Subjects Act (WMO) and did not require full review by an accredited MREC (protocol number 20-606/C).

## Results

### Characteristics of included population

During the 5 days of the study, on average 27.2% (n=970) of the people who visited the testing site could be included; the inclusion was put on hold occasionally during the day due to clogging of testing positions. Of the included people the average age was 42 years (range 18-86 years), the majority was female (n=525, 54.7%) and had symptom onset ≤7 days (n=650, 89.7%). The majority (84.9%) of the samples contained a high viral load (PCR cycle threshold ≤30, E gene copy/ml 2.17E+05) (Table 1). The age and gender distribution of the people included in the study was representative of the tested population in general (average age 38.4 years and 57% female). We did not record reasons for not participating.

**Table 1.** Characteristics of the included population. Data of all included people in the study were analysed by basic demographics, date of onset and PCR results.

|  |  |
| --- | --- |
| **Total (N)** | 970 |
| **Age median (min-max)** | 42 (18-86) |
| **Gender[%F, (n/N)]** | 54.7% (525/960) |
| **Symptoms reported [%Yes, (n/N)]** | 91.3%, 886/970 |
| **Days from symptom onset [median; N]** | 4; 725 |
| **Days 0-3 (n, %)** | 319, 44.0% |
| **Days 4-7 (n, %)** | 331, 45.7% |
| **Days ≥8 (n, %)** | 75, 10.3% |
| **Positivity by PCR [%, (n/N)]** | 19.2% (186/970) |
| **PCR Ct E gene [median (min-max);N]** | 23.6 (15.6--37.4); 186 |
| **Ct ≥35(n, %)** | 1, 0.5% |
| **Ct ≥ 30(n, %)** | 28, 15.1% |
| **Ct ≤ 30(n, %)** | 159, 85.5% |
| **Ct ≤ 25(n, %)** | 113, 60.8% |
| **Ct ≤20(n, %)** | 31, 16.7% |

Abbreviations: n, number; N, total number; F, female; PCR, polymerase chain reaction; E gene, SARS-CoV-2 envelope protein; ct, cycle threshold of PCR

At the time of requesting the appointment, a large majority of the participants had symptoms (91.3%), and most frequently reported cold symptoms and runny nose (64.5%), sore throat (57%), coughing (55%), headache (48%), tiredness (38%), muscle pain (27%), shortness of breath and chills (21%). Some of the more typical and serious symptoms like fever and reproductive cough were only reported by 17% of cases. A very small percentage (1.5%) reported loss of taste and smell.

### Performance of the Ag RDT

The overall sensitivity of the Ag RDT was 84.9% (CI95% 79.1-89.4) and specificity 99.6% (CI95% 98.6-99.9) with a positive and negative predictive value of 98.3% (CI95% 94.0-99.5) and 97.7% (CI95% 96.1-98.7) under an average of 19.2% current prevalence in the region calculated by PCR positivity rate in the test location. Sensitivity improved considerably when analysed by various PCR cycle threshold (ct) intervals showing highest sensitivity for ct values ≤25 (E gene copy/ml 4.87E+06) (99.1% CI95% 95.2-100) and 94.3% (CI95% 89.6-97.0) for ct values ≤30 (E gene copy/ml 2.17E+05). Also sensitivity in people that presented within the first 3 days of symptoms onset was higher than for people later in their disease progression (94.9% vs 90.6%) (Table 2). Overall, the sensitivity was strongly associated with viral load. Three from the four PCR (and culture) negative samples which were detected by the Ag RDT were negative by RT-PCR for other respiratory viruses, while one was weakly positive for rhinovirus (ct>35). Only in one of the samples could metagenomic sequencing identified Rhinovirus B. PCR positive samples which were not detected by the Ag RDT showed a mixed distribution of viral load (n=10/30 <ct 30) and date of onset (5/10 <7 days). For three of these ten no symptoms or contact with a confirmed case was reported and mostly high ct (average ct 33).

**Table 2.** **Sensitivity, specificity, positive and negative predictive values of the antigen RDT compared to RT-PCR stratified by days post onset.** Overall and stratified sensitivity, specificity of Ag RDT was calculated compared to RT-PCR results and days since symptoms onset. Positive and negative predictive values were calculated using 19.2% prevalence setting.

|  | **0-3 days post onset** | **0-7 days post onset** | **All** |
| --- | --- | --- | --- |
| **Clinical Sensitivity (95% CI), N** | 94.9 (86.1-98.3), 319 | 90.6 (84.3-94.6), 650 | 84.9 (79.1-89.4), 970 |
| **Sensitivity ct ≤30, (95% CI), N** | 98.2 (90.6-99.9), 316 | 95.8 (90.5-98.2), 640 | 94.3 (89.6-0.97), 943 |
| **Sensitivity ct ≤25, (95% CI), N** | 100 (92.1-100), 305 | 98.8 (93.7-99.9), 608 | 99.1 (95.2-100), 897 |
| **PPV** | 98.2 (90.7-99.9) | 98.3 (94.0-99.5) | 97.5 (93.8-99.0) |
| **Clinical Specificity (95% CI), N** | 99.6 (97.9-100), 319 | 99.6 (98.6-99.9), 650 | 99.5 (98.7-99.8), 970 |
| **NPV** | 98.9 (96.7-99.6) | 97.7 (96.1-98.7) | 96.5 (0.95-97.6) |

Abbreviations: CI95%, 95% confidence interval; PPV, positive predictive value; NPV, negative predictive value

### Association of Ag RDT results to infectivity

All specimen tested positive by Ag RDT and/or PCR were inoculated on Vero 118 cells. In total 140 from 186 (75%) materials could be cultured and CPE was observed between 2-5 days after inoculation. The culture positive specimen were obtained from individuals with a median of 6 days post onset of disease (range 5-7 days) and high viral load (average ct 25.7, E gene copy/ml 3.15E+06).Mean days since symptom onset was similar for all Ag RDT and/or PCR samples independently of successful culture.

Five from the 140 (3.6%) cultured specimen were Ag RDT negative. These specimen were obtained from slightly older individuals (mean 53, range 35-69), majority female (1/5 male) with a median of 6 days post onset of disease (range 5-7 days, 2 missing) and high viral load (average ct 25.7, E gene copy/ml 3.15E+06). In samples with ct-values <30 (E gene copy/ml 2.17E+05), 6% could not be cultured (10/159) but only 2.5% (4/159) was not detected by Ag RDT. For samples with a ct-value above 30, only 1/27 (4%) could be cultured but 8/27 (30%) were Ag RDT positive indicating that above a ct-value of 30 (E gene copy/ml 2.17E+05) the majority of samples are not infectious which is in agreement with previously published data [10, 11] (Figure 4).

### Significance of time to result

Results were logged at three time points: 5 mins, 10 mins and the recommended readout time of 15 mins and intensity of the test band was recorded. In general, the majority of strong positive samples appeared <5 mins following sample addition (95% of all strong positive results). Bands showing medium intensity had a more equal distribution of time to results in the three time frames while weak positive bands mostly required the recommended 15 mins readout (73% of all weak positive samples) (Figure 3 C). Band intensity correlated directly and strongly with viral load (Figure 3 B).

**Figure 3.**
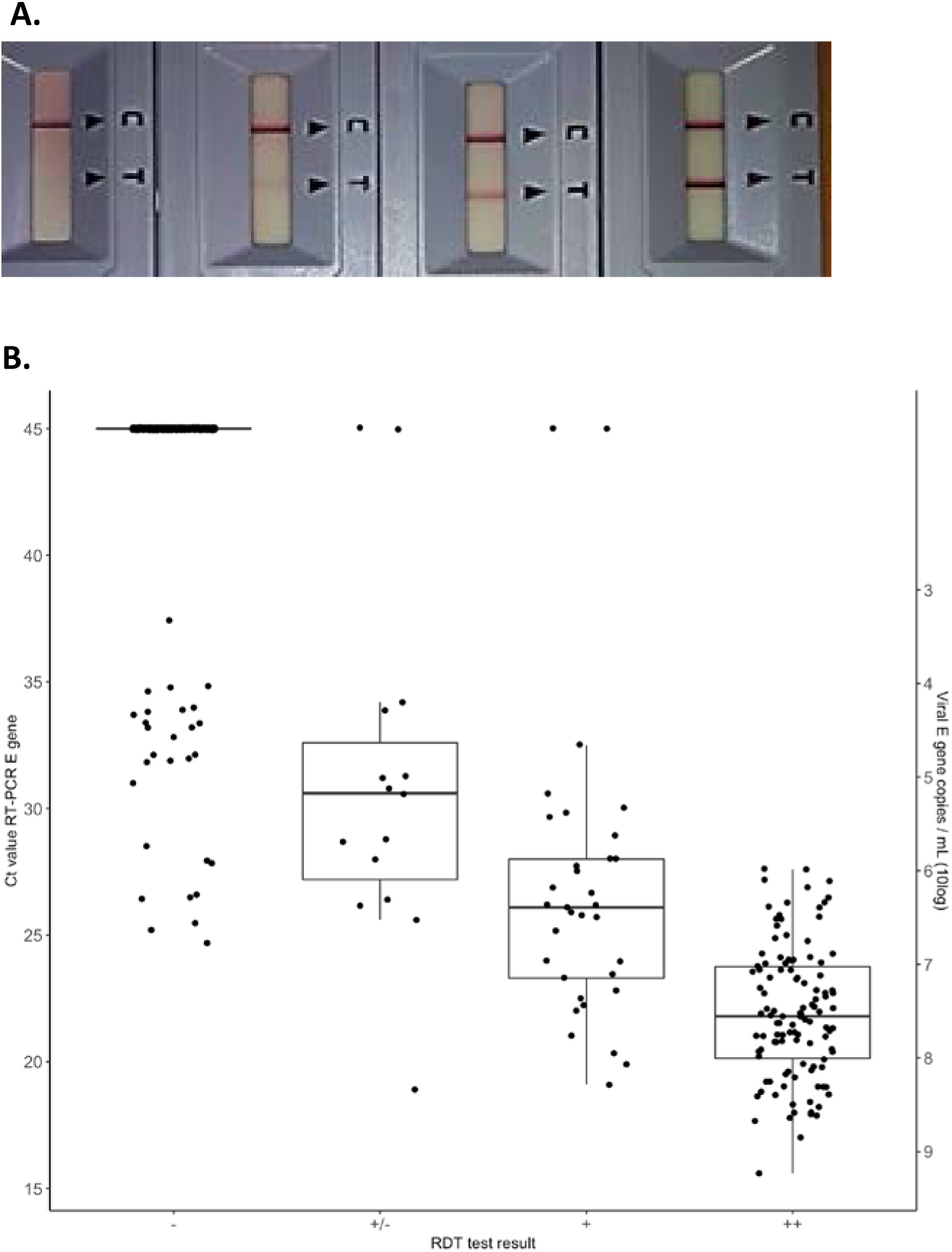
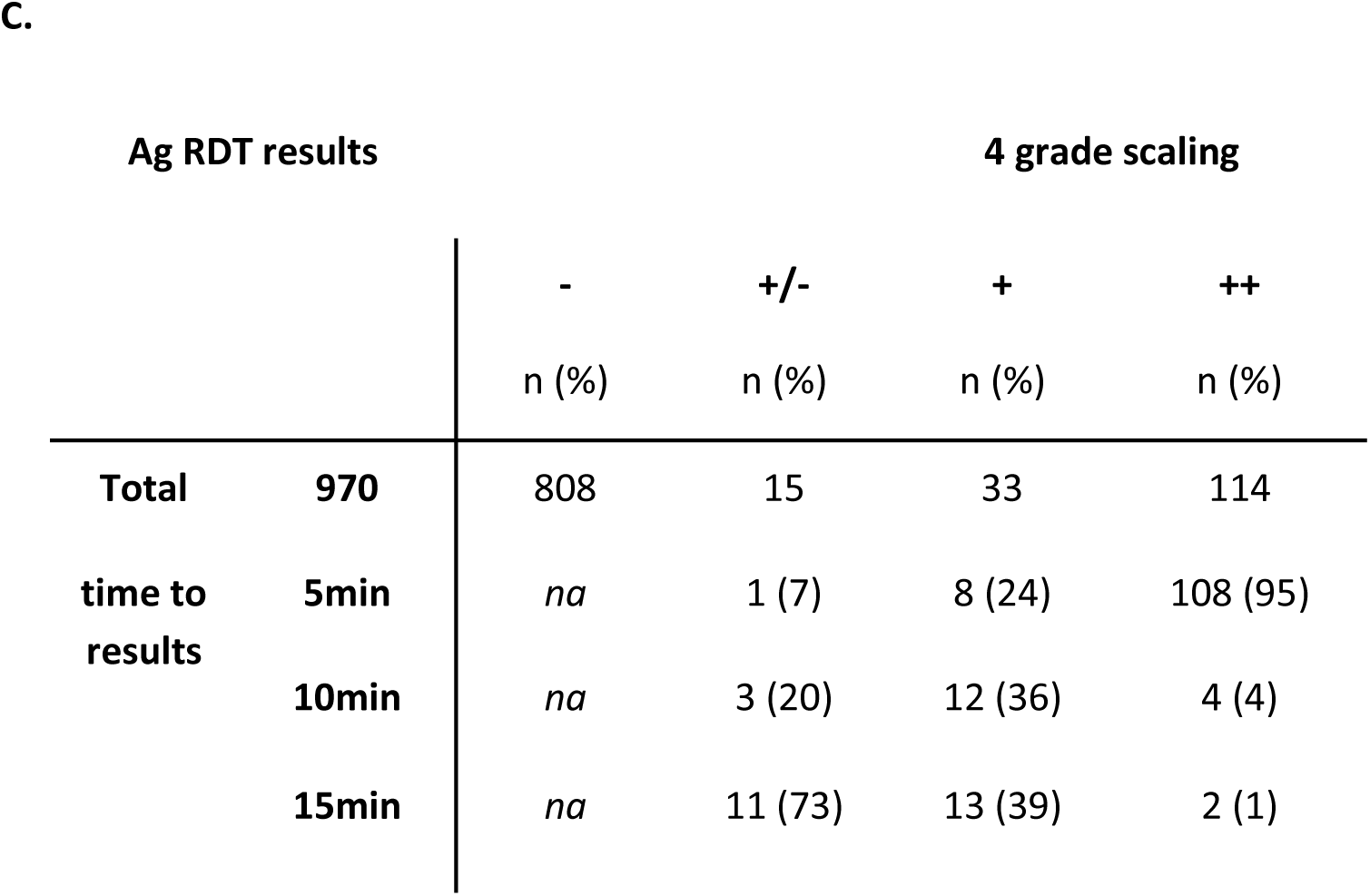
Correlates of time to results. **A**, example of the 4 grade scaling system used; **B**, RT-PCR ct values and viral load correlate well with Ag RDT band intensity (n=970); **C**. Time to results of the tested samples; Abbreviations: ct RT-PCR, cycle threshold reverse transcription polymerase chain reaction; E gene, SARS-CoV-2 envelope protein gene -, negative; +/- weak positive; +, positive; ++, strong positive; n, number; na, not applicable

**Figure 4 A and B.**
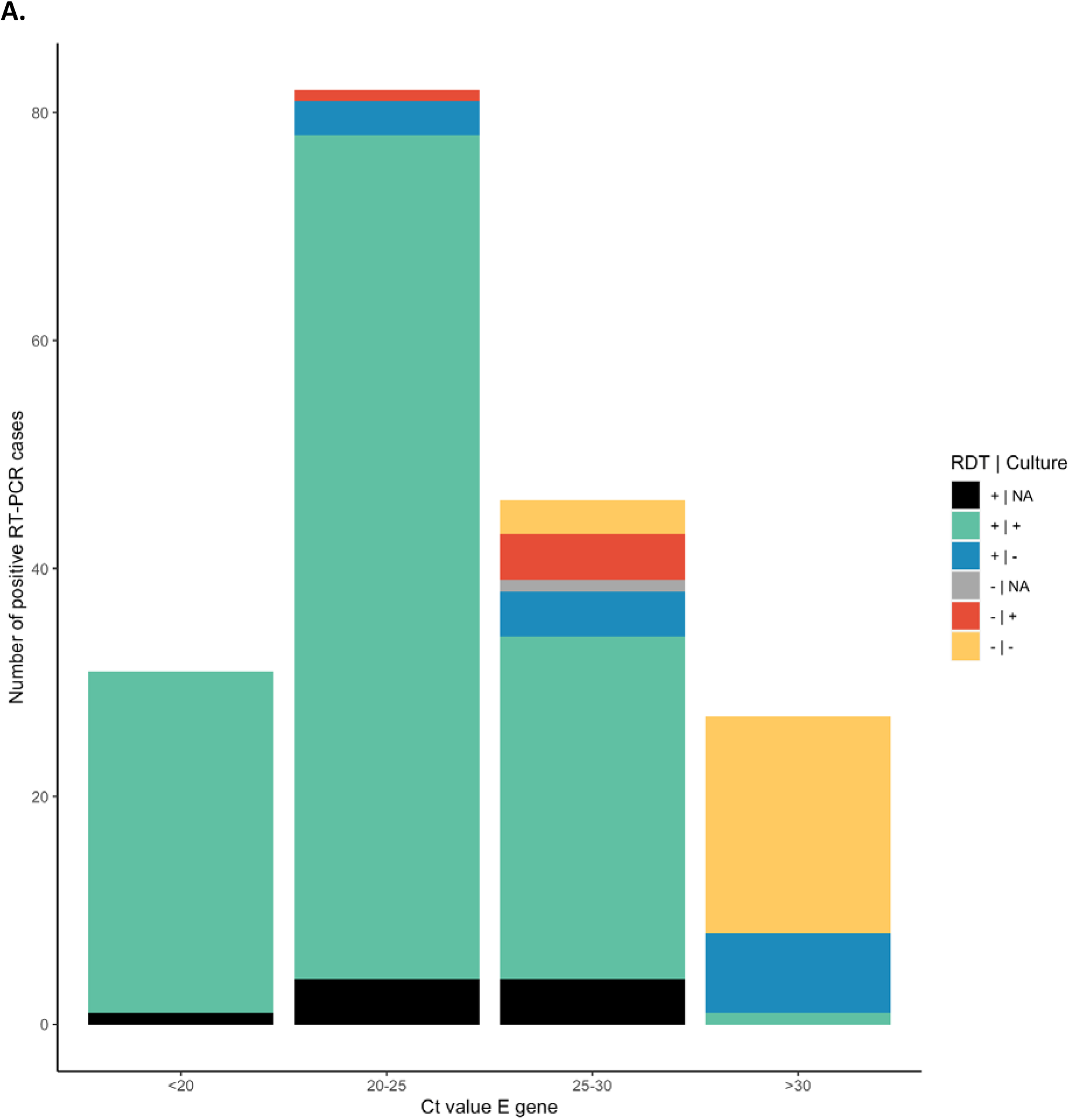
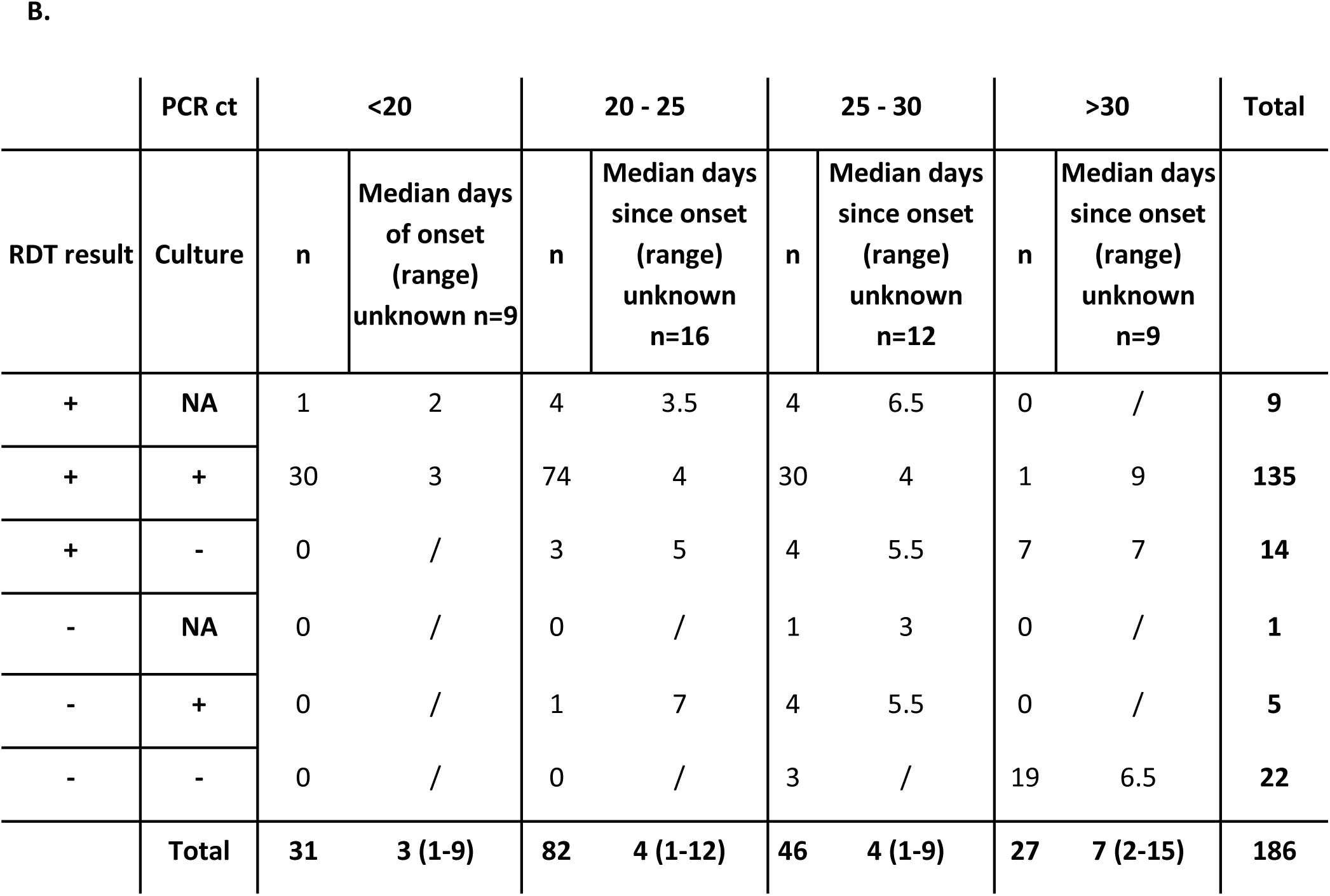
Number of PCR positive samples by Ct value intervals of <20, 20-25, 25-30 and >30 (n=186) in relation to Ag RDT and culture test results. Abbreviations: NA, not applicable ie not cultured; +/- RDT or culture, positive or negative results of the corresponding technique;

## Discussion

Here we describe the results of a large clinical evaluation study using an antigen rapid test under medium-high prevalence setting in a mild symptomatic population to detect SARS-CoV-2 infected people. Overall the test performed well, detecting 84.9% of all cases with RT-PCR as reference. Our results align well with data from other independent evaluations [5]. With this performance the test would fulfil the WHO criteria until the prevalence drops below 2.5% based on positive predictive value. An important question to address is if and how Ag RDT can detect infectious individuals and support the “test, trace and isolate” strategy that is largely employed worldwide to control the COVID-19 pandemic. In our evaluation, we would have detected close to 97% of persons with sufficient viral load to allow virus culture, suggesting screening based on Ag RDT alone in this population would have a high sensitivity for ruling out infectious individuals.

One of the unique strengths of this study is the correlation of results with infectivity. Most PCR positive samples with high viral load could be cultured successfully but a fraction of a potentially infectious group was not detected by the Ag RDT. These patients were in general in the later phase of the infection but still had a high viral load and positive virus cultures. Although, in theory, the presence of antibodies in patients after the first week of onset could reduce the sensitivity of Ag RDT, this does not explain the discrepancy in the samples which were negative in the RDT and positive in the virus culture. We previously demonstrated that the presence of neutralising antibodies do inversely correlate with virus culture [10]. One possible explanation is the use of different samples, causing discrepancy in viral load in the RT-PCR/culture versus Ag RDT samples. However small the proportion, missing infectious people can have serious consequences in certain groups. Testing algorithms should be carefully aligned to these risk/priority groups. On the other hand, Ag RDT could detect cases with relatively low viral load thereby providing a safety margin around the suggested threshold of infectiousness.

Asymptomatic individuals were reported to have similar viral loads to symptomatic people [12] therefore in theory Ag RDT could be used also in this population. This is important as they also contribute to the spread of the virus and not having symptoms might also make people less cautious. Performance data of Ag RDT in this population is however missing and little is known about the viral load of exposed individuals without symptoms. Validation of the Ag RDT test in this situation is recommended, and repeat testing following the calculated incubation time could provide more certainty about the test results.

Currently, there are several Ag RDTs on the market mostly employing NP swabs as a sample. NP and OP swabs are considered the best sample types for detecting SARS-CoV-2 especially in the early phase [2, 12] however the swabbing requires trained personnel and causes discomfort to the patient. Only a few Ag RDTs are marketed directly with a less invasive sample, the nasal swab. Based on the available performance data [13] there is no significant difference in detecting mild symptomatic cases and the use of these more superficially collected nasal swabs seems to be a good alternative. An interesting possibility would be to further explore the use of self-sampling which is one of the potential directions Ag RDT testing will take as it does not require trained personnel, reduces infection risk for the healthcare worker who takes the swab and enables testing for a wider population. Studies indicate somewhat lower efficiency of self-sampling compared to trained professionals further lowering detection rate [14] [15] but evaluation studies are ongoing.

Our study has some limitations. In our setting results of RT-PCR and Ag RDT are compared but in contrast to the instructions for the Ag RDT, two swabs were taken for RT-PCR and virus culture which probably results in a higher amount of viral material collected. This might explain some of the discrepancies between Ag RDT and PCR/culture. Furthermore, the same nostril was used to take the second swab for the Ag RDT which meant to grant comparability between the two tests but might have resulted in lower viral load in the second sample. We used culture as a correlate of infectivity which has certain limitations but it is still the best available technique to measure infectivity. When filling out the questionnaires recall bias by the enrolled people could have affected the data provided. Furthermore, testing is free of charge only for people either with relevant symptoms and/or notified contact with an infected person, therefore some people might have provided symptoms falsely in order to be tested for other reasons.

Despite these possible drawbacks, we conclude that the use of Ag RDT in our drive through test stations would provide a good method to identify the majority of infectious people. For further roll-out, logistics of implementation are important, including a safe working environment for personnel performing the assays if implemented on site, and a system that allows follow-up testing by PCR for risk groups. The national outbreak management team of The Netherlands recommends using Ag RDT for rapid screening, but cautions against sole use of Ag RDTs’ in vulnerable individuals such as persons at risk for severe illness, persons living or working in long term care facilities because of the potential of missed cases. A positive Ag RDT can be used to trigger contact tracing and isolation, but a negative test cannot always rule out infections in these risk populations. A slightly higher risk of missed cases could be taken when persons are screened that have little contact with high risk individuals, although the separation of these is not easy. Ideally, this would be secured through a triage system that guides patients to the proper testing algorithm. In any case, it is imperative to inform people tested by Ag RDT of the potential for false negative tests, and the need for continued behavioural measures.

## Data Availability

all data is available

## Acknowledgement

The mobile lab was kindly provided by the RIVM. Testing was carried out at the Schiedam testing centre where numerous people contributed to the success of this project therefore we would like to thank all involved employees of the GGD Rotterdam - Rijmond and the Schiedam testing centre. We are grateful for the vital involvement of the following volunteers who supported patient inclusion and administrative tasks: Pauline de Best, Loubna Bouzyd, Vera Mols, Jasmijn de Rooij, Louella Kasbergen, Axel Bonacic Marinovic, Maarten Hoek, Kamelia Stanoeva, Nadya Velikova. The SARS-CoV-2 Rapid Antigen Test (Distributed by Roche (SD Biosensor) was provided by the Ministry of Health, Welfare and Sport (VWS). Authors declare no conflict of interest.

## Funding

This work was partly supported by H2020 project RECoVer [grant number 101003589].

## Notes

### Competing Interest Statement

The authors have declared no competing interest.

